# The missing link between genetic association and regulatory function

**DOI:** 10.1101/2021.06.08.21258515

**Authors:** Noah Connally, Sumaiya Nazeen, Daniel Lee, Huwenbo Shi, John Stamatoyannopoulos, Sung Chun, Chris Cotsapas, Christopher A. Cassa, Shamil Sunyaev

## Abstract

The genetic basis of most traits is highly polygenic and dominated by non-coding alleles. It is widely assumed that such alleles exert small regulatory effects on the expression of *cis*-linked genes. However, despite the availability of gene expression and epigenomic data sets, few variant-to-gene links have emerged. It is unclear whether these sparse results are due to limitations in available data and methods, or to deficiencies in the underlying assumed model. To better distinguish between these possibilities, we identified 220 gene-trait pairs in which protein-coding variants influence a complex trait or its Mendelian cognate. Despite the presence of expression quantitative trait loci near most GWAS associations, by applying a gene-based approach we found limited evidence that the baseline expression of trait-related genes explains GWAS associations, whether using colocalization methods (8% of genes implicated), transcription-wide association (2% of genes implicated), or a combination of regulatory annotations and distance (4% of genes implicated). These results contradict the hypothesis that most complex trait-associated variants coincide with homeostatic eQTLs, suggesting that better models are needed. The field must confront this deficit, and pursue this “missing regulation.”

## Introduction

Modern complex trait genetics has uncovered surprises at every turn, including the paucity of associations between traits and coding variants of large effect, and the “mystery of missing heritability,” in which no combination of common and rare variants can explain a large fraction of trait heritability^1^. Further work has revealed unexpectedly high polygenicity for most human traits and very small effect sizes for individual variants. Bulk enrichment analyses have demonstrated that a large fraction of heritability resides in regions with gene regulatory potential, predominantly tissue-specific accessible chromatin and enhancer elements, suggesting that trait-associated variants influence gene regulation^2–4^. Furthermore, genes in trait-associated loci are more likely to have genetic variants that affect their expression levels (expression QTLs, or eQTLs), and the variants with the strongest trait associations are more likely also to be associated with transcript abundance of at least one proximal gene^5^. Combined, these observations have led to the inference that most trait-associated variants are eQTLs, and their effects arise from altering transcript abundance, rather than protein sequence. Equivalent sQTL (splice QTL) analyses of exon usage data have revealed a more modest overlap with trait-associated alleles, suggesting that a fraction of trait-associated variants influence splicing, and hence the relative abundance of different transcript isoforms, rather than overall expression levels. The genetic variant causing expression changes may lie outside the locus and involve a knock-on effect on gene regulation, with the variant altering transcript abundances for genes elsewhere in the genome (a *trans*-eQTL), but the consensus view is that *trans*-eQTLs are typically mediated by the variant influencing a gene in the region (a *cis*-eQTL)^6^. Thus, a model has emerged in which most trait-associated variants influence proximal gene regulation.

Here we argue that this unembellished model—in which GWAS peaks are mediated by the effects on the homeostatic expression in bulk tissues—is the exception rather than the rule. We highlight challenges of current strategies linking GWAS variants to genes and call for a reevaluation of the basic model in favor of more complex models possibly involving context-specificity with respect to cell types, developmental stages, cell states, or the constanstancy of expression effects.

Our argument begins with several observations that challenge the unembellished model. One challenge is the difference between spatial distributions of eQTLs, which are dramatically enriched in close proximity to genes, and GWAS peaks, which are usually farther away^7–9^. Another is that expression levels mediate a minority of complex trait heritability^10^. Finally, many studies have designed tools for colocalization analysis: a test of whether GWAS and eQTL associations are due to the same set of variants, not merely distinct variants in linkage disequilibrium. If the model is correct, most trait associations should also be eQTLs, but across studies, only 5-40% of trait associations co-localize with eQTLs^11–14^.

Despite the doubts raised, the fact that most GWAS peaks do not colocalize with eQTLs cannot disprove the predominant, unembellished model. In a sense, negative colocalization results are confusing because their hypothesis is too broad. If we predict merely that GWAS peaks will colocalize with *some* genes’ expression, it is not clear what is meant by a peak’s failure to colocalize with *any individual* gene’s expression.

Thus, a narrower, more testable hypothesis requires identifying genes we believe *a priori* are biologically relevant to the GWAS trait. If these trait-linked genes have nearby GWAS peaks and eQTLs, failure to colocalize would be a meaningful negative result.

Earlier studies tested all GWAS peaks; when a peak has no colocalization, the model is inconclusive. But trait-linked genes that fail to colocalize reveal that our method for detecting non-coding variation is, with current data, incompatible with our model for understanding it.

With this distinction in mind, we created a set of trait-associated genes capable of supporting or contradicting the model of non-coding GWAS associations acting as eQTLs. For this purpose the selection of genes becomes extremely important. Because the model attempts to explain the genetic relationship between traits and gene expression, true positives cannot be selected based on measurements of genetic association to traits (GWAS) or expression (eQTL mapping). With this restriction, one source of true positives is to identify genes that are both in loci associated with a complex trait and are also known to harbor coding mutations tied to a related Mendelian trait or the same complex trait. Using a model not based on expression, Mendelian genes are enriched in common-variant heritability for cognate complex traits^15^. The genes and their coding variants may be detected in familial studies of cognate Mendelian disorders, or by aggregation in a burden test on the same complex phenotypes as GWAS^16^

For genes whose coding variants can cause detectable phenotypic change, the strong expectation is that a variant of small effect influences the gene identified by its rare coding variants. As an example, *APOE* and *LDLR* are both low-density lipoprotein receptor genes^17,18^. Coding variants in *APOE* and *LDLR* can lead to the Mendelian disorder familial hypercholesterolemia^18,19^. Even in the absence of a Mendelian coding variant, experiments in animal models have found that the overexpression of these genes reduces cholesterol levels^20–22^. GWAS on human subjects have found significant associations near *APOE* and *LDLR*, so it seems reasonable to suspect that any noncoding effects in these loci may be mediated by these genes. This general relationship between Mendelian and complex traits is supported by several lines of evidence summarized in **Supplementary Note 1**.

## Results

To test the model that trait-associated variants influence baseline gene expression, we assembled a list of such putatively causative genes. We selected seven polygenic common traits with available large-scale GWAS data, each of which also has an extreme form in which coding variants of large effect alter one or more genes with well-characterized biology (**Table 1**). Our selection included four common diseases: type II diabetes^23^, where early onset familial forms are caused by rare coding mutations (insulin-independent MODY; neonatal diabetes; maternally inherited diabetes and deafness; familial partial lipodystrophy); ulcerative colitis and Crohn disease^24,25^, which have Mendelian pediatric forms characterized by severity of presentation; and breast cancer^26^, where germline coding mutations (e.g., *BRCA1*) or somatic tissue (e.g., *PIK3CA*) are sufficient for disease. We also chose three quantitative traits: low and high density lipoprotein levels (LDL and HDL); and height.

**Table 1.** Putatively causative Mendelian genes. Each gene includes reference(s) to the known biological role of its coding variants, as established in familial studies, *in vitro* experiments, and/or animal models. Genes from Backman *et al*. are not included here, but can be found in **Figure 3**.

| Phenotype | Genes |
| --- | --- |
| LDL | APOB<sup>46,47</sup><br>APOC2<sup>48</sup><br>APOE<sup>49</sup><br>LDLR<sup>50</sup><br>LPL<sup>51,52</sup><br>PCSK9<sup>53</sup> |
| HDL | ABCA1<sup>54-56</sup><br>APOA1<sup>57</sup><br>CETP<sup>58</sup><br>LIPC<sup>59-61</sup><br>LIPG<sup>62</sup><br>PLTP<sup>63</sup><br>SCARB1<sup>64,65</sup> |
| Height | ANTXR1<sup>66,67</sup><br>ATR<sup>68,69</sup><br>BLM<sup>70,71</sup><br>CDC6<sup>72</sup><br>CDT1<sup>72,73</sup><br>CENPJ<sup>74</sup><br>COL1A1<sup>75</sup><br>COL1A2<sup>76,77</sup><br>COMP<sup>78,79</sup><br>CREBBP<sup>80-82</sup><br>DNA2<sup>83</sup><br>EP300<sup>84,85</sup><br>EVC<sup>86,87</sup><br>EVC2<sup>87,88</sup><br>FBN1<sup>89-92</sup><br>FGFR3<sup>93-95</sup><br>FKBP10<sup>96-98</sup><br>GHR<sup>99-102</sup><br>KRAS<sup>103-105</sup><br>NBN<sup>106,107</sup><br>NIPBL<sup>108,109</sup><br>ORC1<sup>72,73,110</sup><br>ORC4<sup>72,73</sup><br>ORC6L<sup>72,111</sup><br>PCNT<sup>112-114</sup><br>PLOD2<sup>115-117</sup><br>PTPN11<sup>118-120</sup><br>RAD21<sup>121-123</sup><br>RAF1<sup>124,125</sup><br>RECQL4<sup>126-128</sup><br>RIT1<sup>129-131</sup><br>ROR2<sup>132-134</sup><br>SLC26A2<sup>135-137</sup><br>SMAD4<sup>138-140</sup><br>SRCAP<sup>141,142</sup><br>WRN<sup>143-145</sup> |
| Crohn disease | ATG16L1<sup>146</sup> |
|  | CARD9<sup>147</sup><br>IL10<sup>148</sup><br>IL10RA<sup>149,150</sup><br>IL10RB<sup>151,152</sup><br>IL23R<sup>153–155</sup><br>IRGM<sup>156–158</sup><br>NOD2<sup>159,160</sup><br>PRDM1<sup>161</sup><br>PTPN22<sup>162</sup><br>RNF186 |
| Ulcerative colitis | ATG16L1<sup>163</sup><br>CARD9<sup>147</sup><br>IL23R<sup>155,164</sup><br>IRGM<sup>156</sup><br>PRDM1<sup>161</sup><br>PTPN22<sup>162</sup><br>RNF186<sup>165,166</sup> |
| Type II diabetes | ABCC8<sup>167</sup><br>BLK<sup>168</sup><br>CEL<sup>169,170</sup><br>EIF2AK3<sup>171–173</sup><br>GATA4<sup>174</sup><br>GATA6<sup>175,176</sup><br>GCK<sup>177</sup><br>GLIS3<sup>178</sup><br>HNF1A<sup>179,180</sup><br>HNF1B<sup>181,182</sup><br>HNF4A<sup>183,184</sup><br>IER3IP1<sup>185–187</sup><br>INS<sup>188</sup><br>KCNJ11<sup>189,190</sup><br>KLF11<sup>191</sup><br>LMNA<sup>192</sup><br>NEUROD1<sup>193</sup><br>NEUROG3<sup>194–196</sup><br>PAX4<sup>197–199</sup><br>PDX1<sup>200–202</sup><br>PPARG<sup>203,204</sup><br>PTF1A<sup>205</sup><br>RFX6<sup>206,207</sup><br>SLC19A2<sup>208–210</sup><br>SLC2A2<sup>211,212</sup><br>WFS1<sup>213–215</sup><br>ZFP57<sup>216,217</sup> |
| Breast cancer (selected using MutPanning<sup>218</sup>) | AKT1<br>ARID1A<br>ATM<br>BRCA1<br>BRCA2<br>CBFB<br>CDH1<br>CDKN1B |
|  | CHEK2<br>CTCF<br>ERBB2<br>ESR1<br>FGFR2<br>FOXA1<br>GATA3<br>GPS2<br>HS6ST1<br>KMT2C<br>KRAS<br>LRRC37A3<br>MAP2K4<br>MAP3K1<br>NCOR1<br>NF1<br>NUP93<br>PALB2<br>PIK3CA<br>PTEN<br>RB1<br>RUNX1<br>SF3B1<br>STK11<br>TBX3<br>TP53<br>ZFP36L1 |

In well-powered GWAS, even relatively rare large-effect coding alleles (mutations in *BRCA1* which cause breast cancer, for instance) may be detectable as an association to common variants, which could make the effect of a coding variant appear to be regulatory instead. To account for this possibility, we computed association statistics in each GWAS locus conditional on coding variants. We applied a direct conditional test to datasets with available individual-level genotype data (height, LDL, HDL); for those studies without available genotype data, we computed conditional associations from summary statistics using COJO^27,28^ (**Methods**). With both methods, the resulting GWAS associations should reflect only non-coding variants.

After controlling for coding variation, we examined whether these genes are more likely than chance to be in close proximity to variants associated with the polygenic form of each trait. In agreement with existing literature^29^, we observe a significant enrichment for all traits in our combined Mendelian and Backman *et al*. gene sets (**Supp. Fig. 1**).

Of our 220 genes, 147 (67%) fell within 1 Mb of a GWAS locus for the cognate complex trait, over three times as many as the 43 predicted by a random null model (95% confidence interval: 31.5-54.5). Our window of 1 Mb represents roughly the upper bound for distances identified between enhancer-promoter pairs, but most pairs are closer^30^, so we would expect enrichment to increase as the window around genes decreases; this proves to be the case. At a distance of 100 kb, we find 104 putatively causative genes (47%), though the null model predicts only 11 (95% CI 4.5-17.0), a order-of-magnitude enrichment (**Supp. Fig. 1**). Given their known causal roles in the severe forms of each phenotype, these results suggest that the 147 genes near GWAS signals are likely to be the targets of trait-associated non-coding variants. For example, we see a significant GWAS association between breast cancer risk and variants in the estrogen receptor (*ESR1*) locus even after controlling for coding variation; the baseline expression model would thus predict that non-coding risk alleles alter *ESR1* expression to drive breast cancer risk.

We next looked for evidence that the trait-associated variants were also altering the expression of our 147 genes in relevant tissues. Controlling for the number of tests we conducted, 134 of these genes had an eQTL in at least one relevant tissue at a false discovery rate of Q < 0.05 (**Methods**). If these variants act through changes in gene expression, phenotypic associations should be driven by the same variants as eQTLs in relevant tissue types. We therefore looked for co-localization between our GWAS signals and eQTLs in relevant tissues (**Table 2**) drawn from the GTEx Project, using three well-documented methods: coloc^11^, JLIM^12^, and eCAVIAR^14^. We found support for the colocalization of trait and eQTL association for only 7 genes out of 147 (4.8%) for coloc; 10/147 (6.8%) for JLIM; and 8/147 (5.4%) for eCAVIAR. Accounting for overlap, this represents only 18/220 putatively causative genes (8.2%) or 18/147 (12.2%) putatively causative genes near GWAS peaks, even without full multiple-hypothesis testing correction (**Methods**), which is not obviously better than random chance. We note that prior estimates of the fraction of *GWAS associations* colocalizing with eQTLs (25%-40%^11,12,14,31^) do not directly evaluate the ability to find causative genes. By contrast, our estimate of the number of putatively causative *genes* that colocalize with eQTLs tests the consistency of our knowledge, models, and data.

**Table 2.** Tissue-trait pairs. Tissues were selected for each trait based on *a priori* knowledge of disease biology.

| Mendelian trait | GWAS trait | Tissues examined |
| --- | --- | --- |
| Breast cancer | Breast cancer | Breast mammary tissue |
| Crohn disease | Crohn disease | Small intestine terminal ileum<br>Colon Sigmoid<br>Colon Transverse |
| Ulcerative colitis | Ulcerative colitis | Small intestine terminal ileum<br>Colon Sigmoid<br>Colon Transverse |
| Dyslipidemia<br>Hyperlipidemia<br>Tangier's disease | HDL | Liver<br>Adipose (subcutaneous)<br>Whole blood |
| Dyslipidemia<br>Hyperlipidemia | LDL | Liver<br>Adipose (subcutaneous)<br>Whole blood |
| Mendelian short stature | Height | Skeletal muscle |
| Monogenic diabetes | Type II diabetes | Pancreas<br>Skeletal muscle<br>Adipose (subcutaneous)<br>Small intestine terminal ileum |

A potential weakness of our approach is the restriction of our search to pre-defined tissues. We believe this is necessary in order to avoid the disadvantages of testing each gene-trait pair in each tissue—either a large number of false positives, or a severe multiple-testing correction that may lead to false negatives. However, restricting to the set of tissues with a known biological role and available expression data almost certainly leaves out tissues with relevance in certain contexts. Some of the tissues we do use have smaller sample sizes, limiting their power to detect eQTLs with smaller effects.

To address potential shortcomings from the available sample of tissue contexts, we incorporated the Multivariate Adaptive Shrinkage Method (MASH)^32^. MASH is a Bayesian method that takes genetic association summary statistics measured across a variety of conditions and, by determining patterns of similarity across conditions, updates the summary statistics of each individual condition. In our case, if an eQTL is difficult to find in a tissue of interest, incorporating information from other tissues may help us detect it. Unlike meta-analysis, this method generates summary statistics that still correspond to a specific tissue.

We ran MASH on every locus used in our earlier analysis, using data from all non-brain GTEx tissues (**Methods**). Rerunning coloc with these modified statistics increased the number of GWAS-eQTL colocations from 389 to 489. However the 100 new colocations identified only four additional putatively causative genes (**Supp. Fig. 2**). These results indicate that tissue type selection was not the limiting factor in our analysis.

Transcriptome-wide association studies (TWAS)^33–36^ are another class of methods applied to identify causative genes under GWAS peaks using gene expression. TWAS measures genetic correlation between traits and is not designed to avoid correlations caused by LD, which gives it higher power in the case of allelic heterogeneity or poorly typed causative variants^37^. However, while sensitive, TWAS analyses typically yield expansive result sets that include many false positives and are sensitive to the number of tissue types^37^. Results from the FUSION implementation of TWAS^35^ across all tissues identified our putatively causative genes as likely tied to the GWAS peak in 66/220 loci (30%). However, only 4/220 (1.8%) genes were identified by FUSION when we restricted the analysis to relevant tissues.

Given the paucity of expression-mediated GWAS peaks, we asked whether GWAS variants indeed reside in regulatory sites. Taking the 128 genes in the Mendelian subset of putatively causative genes, we fine-mapped each nearby GWAS association using the SuSiE algorithm^38^. For 37 of these genes, we identified at least one high-confidence fine-mapped SNP (PIP>0.7) within 100kb of the transcription start site. We tested whether these fine-mapped SNPs fall within regulatory DNA marked by chromatin accessibility^39^, a narrowly mapped active histone modification feature (H3K27ac, H3K4me1, or H3K4me3^40^), or characterized as an “enhancer” by ChromHMM^41,42^ (**Methods**). As many as 32/37 (86%) genes identified this way have a fine-mapped SNP within a regulatory feature across all the tissue types examined, or 25/37 (68%) when restricting to phenotype-relevant tissues (**Fig. 3**; **Supp. Table 1, Supp. Table 2**). Despite strong evidence that these GWAS associations are due to regulatory effects, only 5/25 loci (20%) demonstrably correspond to expression effects in our eQTL analysis.

**Figure 1.**
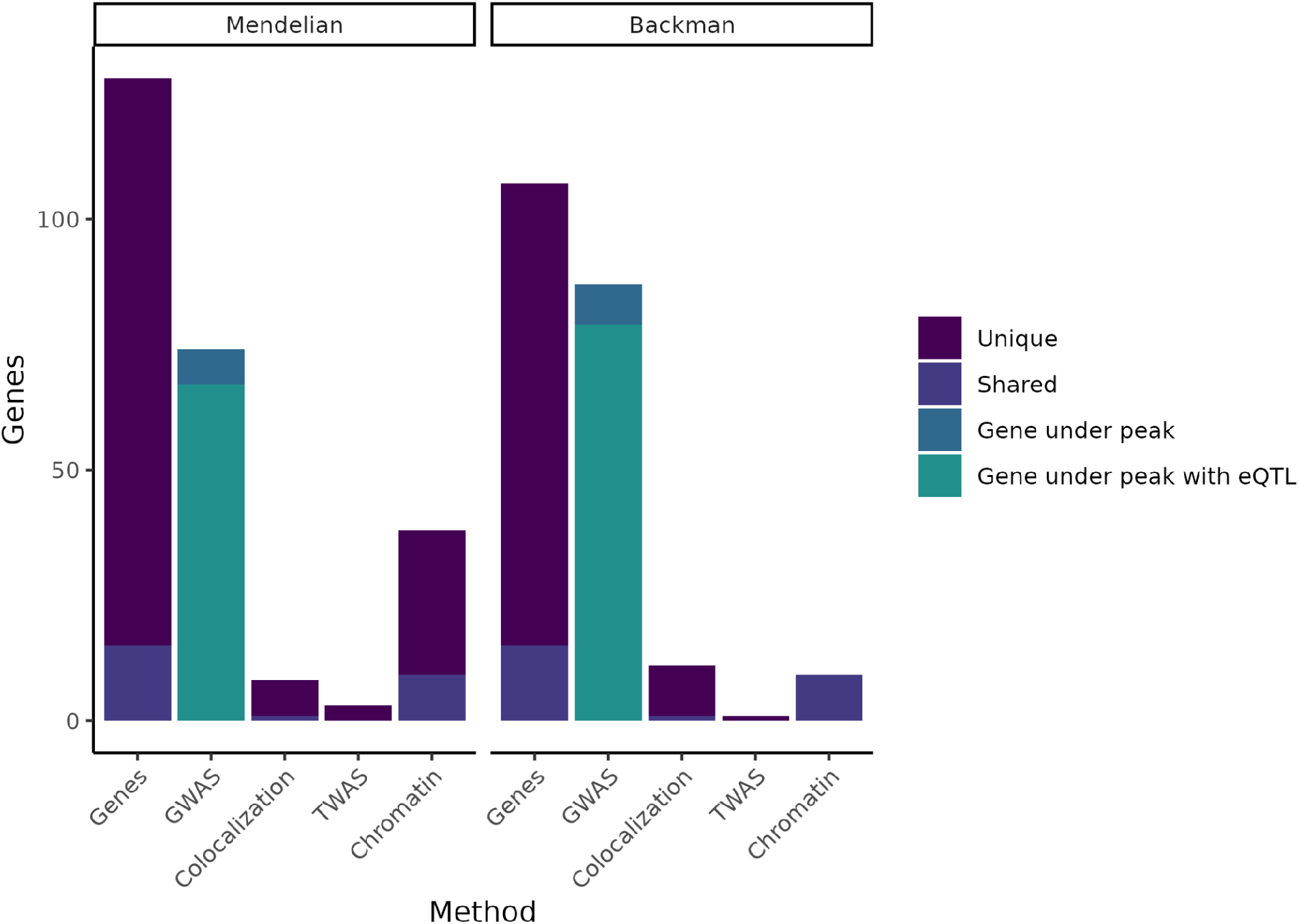
Putatively causative genes identified by each method category. The leftmost column in each half of the plot displays the entire group of putatively causative genes for our Mendelian set of genes and our Backman *et al*. set of genes respectively, as well as noting how many are unique to each set or shared between the two sets. The second column in each half indicates how many genes from each set have a nearby GWAS peak, or have both a nearby GWAS peak and an eQTL. The remaining columns indicate how many genes were identified through colocalization, TWAS, or chromatin methods, while noting how many of these genes are unique vs. shared between the Mendelian and Backman sets.

**Figure 2.**
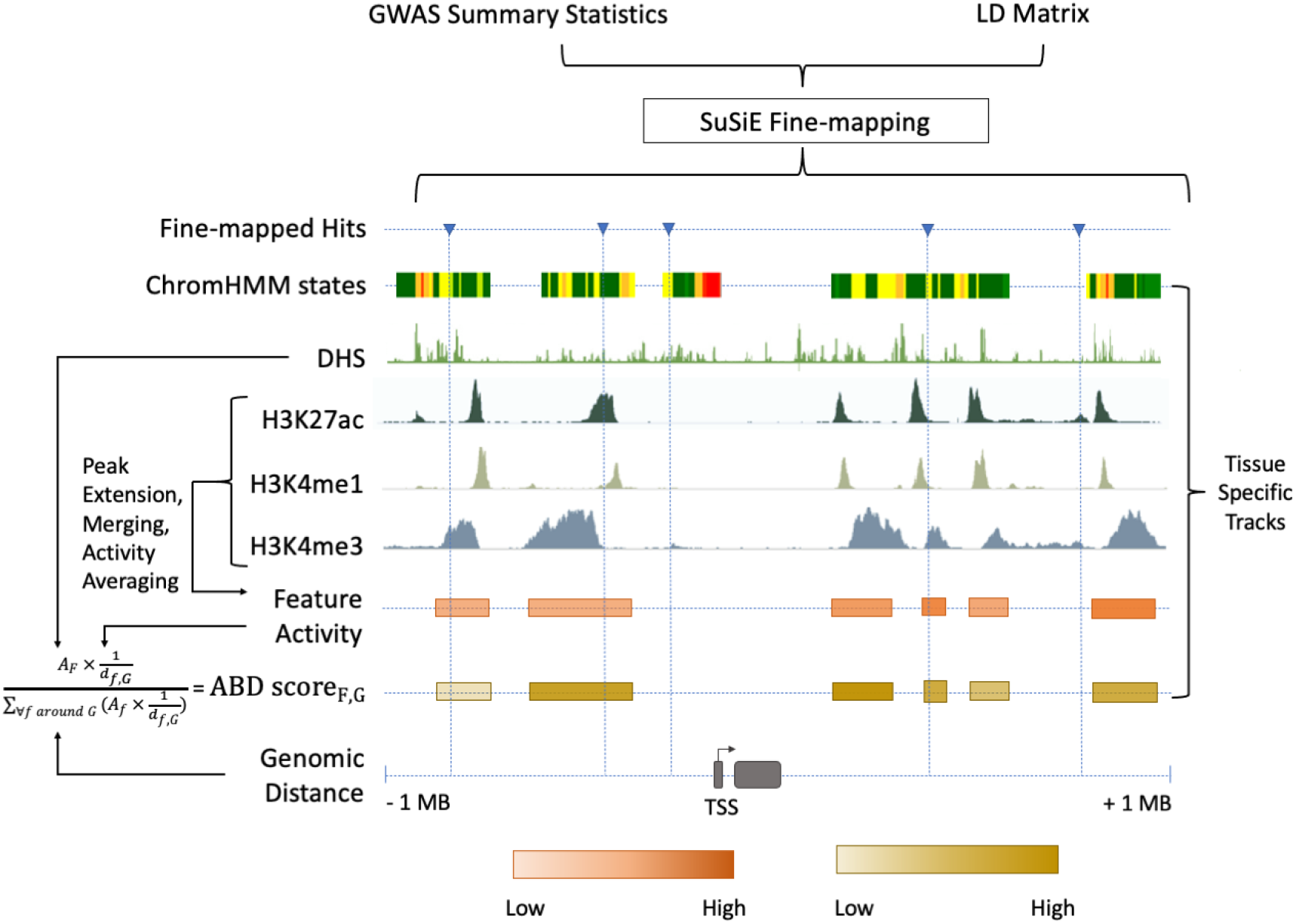
Chromatin-based causative gene identification. Following the fine-mapping of GWAS variants, three parallel methods were used. The first identified fine-mapped variants falling within regions annotated as enhancers by ChromHMM. The second identified variants within histone modification features, and evaluated their relevance using an ABD score that combined the strength of the feature (i.e., the strength of the acetylation or methylation peak) with its genomic distance to the gene of interest (**Methods**). The third repeated both of these—checking for fine-mapped variants within a region and calculating the ABD score—for DNase I hypersensitivity sites.

**Figure 3.**
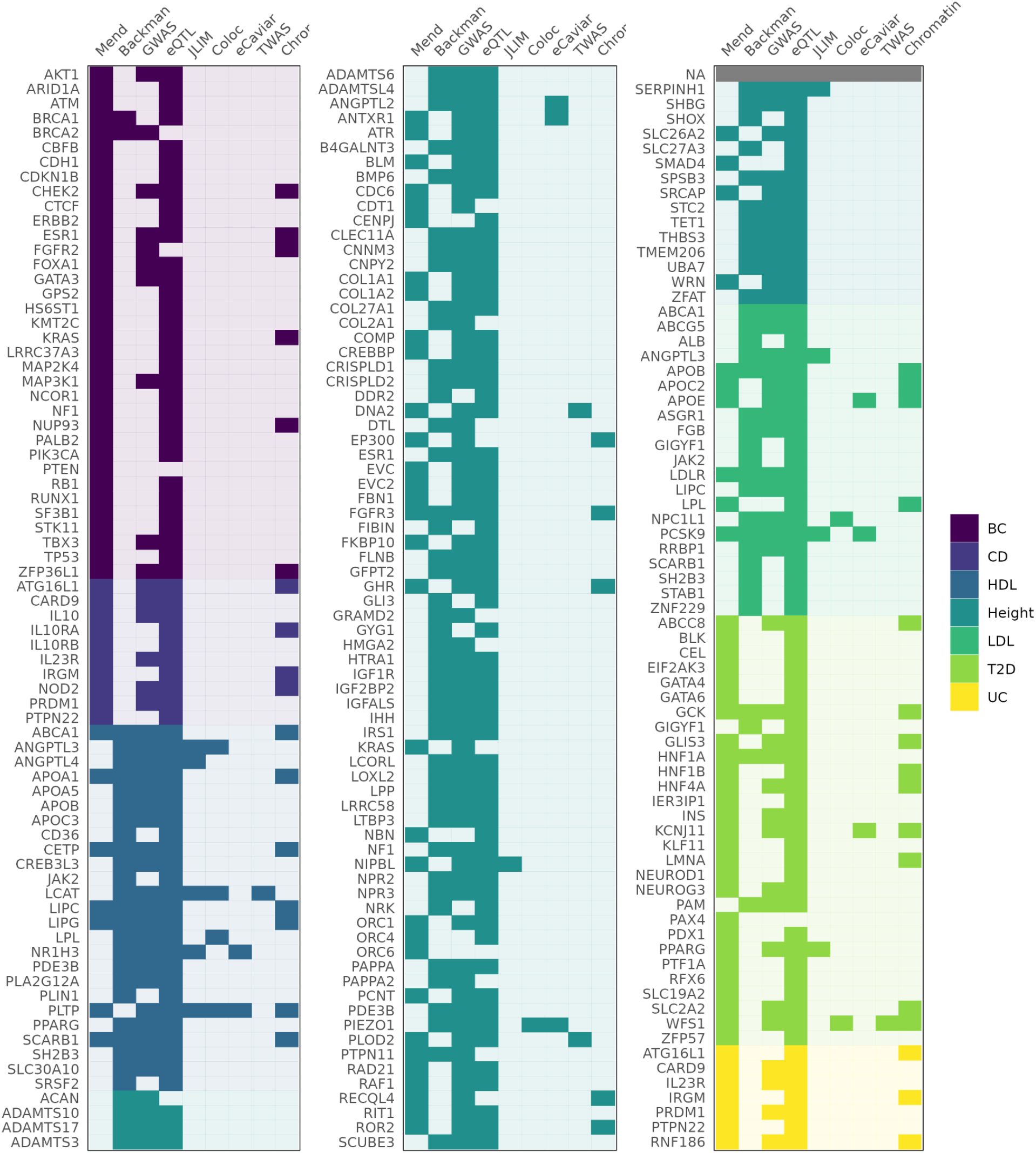
Genes identified as associated with a complex trait by each method. Columns “Mend” and “Backman” indicate whether a gene is from the Mendelian set of putatively causative genes, the Backman et al. set, or both. Subsequent columns indicate whether a gene was identified as a hit using each of our methods: JLIM, coloc, eCaviar, TWAS, and chromatin analysis.

In order to more directly compare our regulatory feature analysis to our eQTL analysis, we measured “activity-by-distance”—a simplification of the “activity-by-contact” method^30,43^ (**Methods; Fig. 2**). Taking each locus’s feature with the highest ABD score, we implicate 5/37 (14%) of our Mendelian subset of genes. This reinforces our observation that, even when a GWAS association and trait-relevant gene are proximal, they are difficult to link, whether using eQTLs or chromatin data.

## Discussion

Overall, our results are consistent with the idea that complex traits are governed by non-coding genetic variants whose effects on phenotype are mediated by their contribution to the regulation of nearby genes. However, these results are inconsistent with the model that a common mechanism of this mediation is the effect on baseline expression within tissues. The enrichment of our putatively causative genes—selected based on existing biological knowledge—near GWAS peaks supports their role in complex traits. Additionally, the enrichment of fine-mapped GWAS variants in accessible chromatin regions and regulatory features lends support to the model of GWAS associations being produced by eQTLs. However, the inability of varied statistical methods to actually link GWAS associations and expression contradicts the idea that the causative GWAS variants are homeostatic, bulk-tissue eQTLs of the sort found in broad expression-data collection projects.

Many explanations have been suggested for the limited success of expression methods to explain the mechanisms of GWAS variants. Undirected, broad approaches— including most GWAS-eQTL linking studies—are designed to be largely independent of *a priori* biological knowledge and hypotheses. This unconstrained focus is ideal for discovery, but, though it delivers the largest number of positive findings, it is ill-suited to provide an explanation for negative results—when you don’t know what you were looking for, it’s hard to explain why you didn’t find it. By testing only loci for which there is a strongly suspected contributing gene, we are better able to distinguish which factors prevent us from identifying it using expression.

As a result, we conclude that a number of explanations often considered when evaluating expression-based variant-to-gene methods are not applicable in the context we examined. These include non-expression-mediated mechanisms, lack of statistical power for GWAS, the absence of eQTLs for relevant genes, and underpowered methods for linking expression to GWAS (**Table 3**).

**Table 3.**
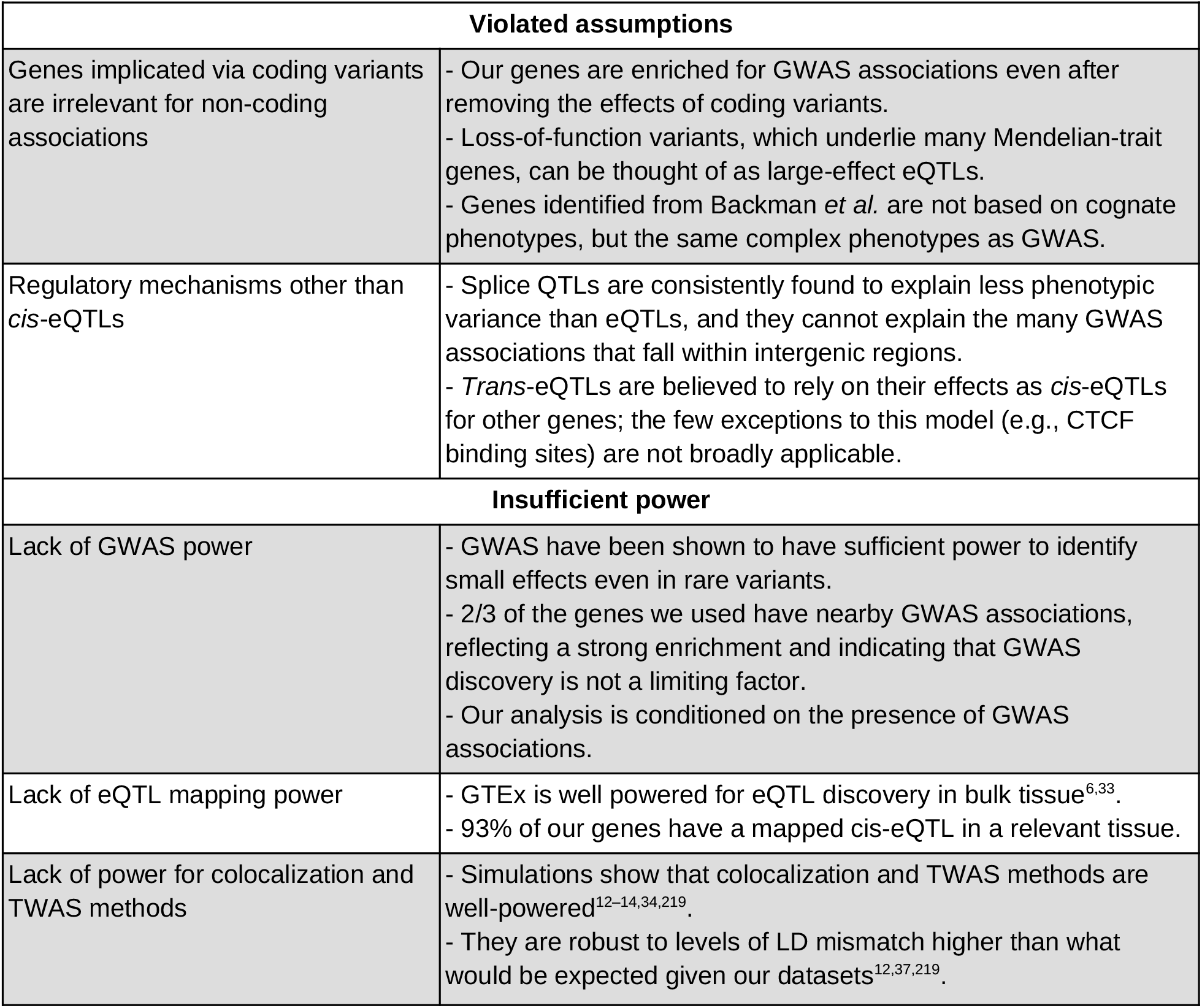
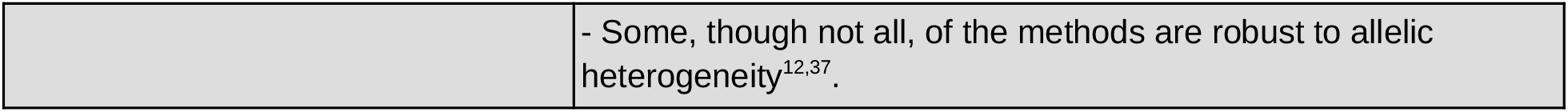
Proposed explanations for negative results under the unembellished model. Many explanations have been proposed for GWAS associations that are not explained by *cis*-eQTLs. This table details explanations inconsistent with our results, which are explained in the left column and addressed in the right. Explanations involving more detailed models of gene regulation can be found in **Table 4**. Two of the explanations addressed here involve violations of the assumptions of our and other expression-based complex trait studies. If coding and non-coding variants affect fundamentally different biological pathways, or if trait associations rarely depend on *cis*-eQTLs, our methods of mapping regulation to traits would have nothing to uncover. Even in the presence of eQTL-driven trait associations, insufficient power to detect trait associations, to detect eQTL associations, or to link the two would result in predominantly negative results.

Instead, we believe the “missing regulation” will be found primarily through examining more nuanced models of gene expression. Solving the mystery will require not only identifying the eQTLs behind GWAS peaks, but also explaining the phenotypic irrelevance of our “red herring eQTLs”: eQTLs for putatively causative genes that fall near GWAS peaks but do not colocalize with them. Some proposed models involve expression that depends on context—whether cell type, cell state, environment, or developmental stage. Others depend on heterogeneity of expression or the variance of expression across relatively short time scales. These various models may depend on or be augmented by thresholding or buffering—processes causing a change in gene expression to have a non-linear effect on phenotype. A summary of these models can be found in **Table 4**.

**Table 4.** Explaining negative results with more nuanced models of gene regulation. To reconcile an expression-based model with our observations requires us to both explain the absence of trait-linked eQTLs as well as explaining away the inconsequence of eQTLs for trait-linked genes. The left-hand side lists additions or changes to the unembellished model, while the right contains explanations of the models and current relevant research.

| Extended models of gene regulation |  |
| --- | --- |
| Context dependency:<br>A context-specific eQTL, invisible in bulk tissue, replaces or supplements the bulk tissue homeostatic eQTL | Cell type<sup>220–234</sup><br>- Only a subset of cell types in the tissue contribute to the GWAS phenotype.<br>- An eQTL specific to such a cell type is causative for the phenotype.<br>- The eQTL cannot be detected in bulk tissue because of the cell type's low prevalence. |
|  | Developmental timing<sup>220,235–240</sup><br>- The GWAS phenotype is influenced by an earlier point in individual development or cell differentiation.<br>- eQTLs present at the correct moment contribute to phenotype, but eQTLs observed later do not. |
|  | Cell state or environment<sup>225,227,231,233,241–248</sup><br>- The causative eQTL has effects that are undetectable in steady-state expression under normal conditions.<br>- It may activate only in response to a specific environmental condition, such as infection.<br>- It may depend on the stage of the cell cycle. |
| Non-linear or non-homeostatic:<br>The relationship between eQTL and genotype is indirect | Nonlinearity<sup>249–269</sup><br>- There may be buffering that prevents a change in expression from producing a change in protein levels.<br>- Expression below a certain level may not influence phenotype, rendering small eQTLs irrelevant. |
|  | Steady-state expression may be a poor model<sup>270–281</sup><br>- Phenotype may depend on the kinetics of expression, which |
|  | could be cyclical or follow some other pattern.<br>- Expression may be stochastic, such that only a random subset of cells display the relevant expression pattern at any one time. |

The implications of our results are both conceptual and practical. The inability of current models to identify the expression effects of known trait-associated genes, and to explain the non-effects of their identified eQTLs, calls for new models of the role of gene regulation in complex traits. One long-standing goal of GWAS has been to discover genes contributing to complex traits^1,8^, but low rates of positive findings for expression-based variant-to-gene methods have constrained this possibility^12,44^. Among other challenges, this has limited the benefit of GWAS and expression data for disease-gene mapping and drug discovery^44,45^. Another practical question raised is the value of different large-scale datasets. Compared to genotypes, expression data are relatively difficult to collect. If the most relevant models are shown to depend on effects not observable in bulk-tissue, homeostatic eQTL mapping, the field may need to consider prioritizing other forms of expression data.

The introduction to this manuscript includes two examples of suspect genes: *APOE* and *LDLR*. Both genes harbor coding variants causing Mendelian hypercholesterolemia. Both have non-coding variants that GWAS have tied to LDL levels. Both have eQTLs in trait-relevant tissues. For *APOE*, these points cohere into an explanation: the LDL-association is an eQTL for the lipid-binding gene. But for *LDLR*—and for most genes— the association, the mechanism, and the gene cannot be tied together. In the field of complex-trait genetics—both basic and translational—solving this regulatory mystery may prove to be a critical step.

## Data Availability

All relevant data are included within the paper.

## Figures

**Supplementary figure 1.**
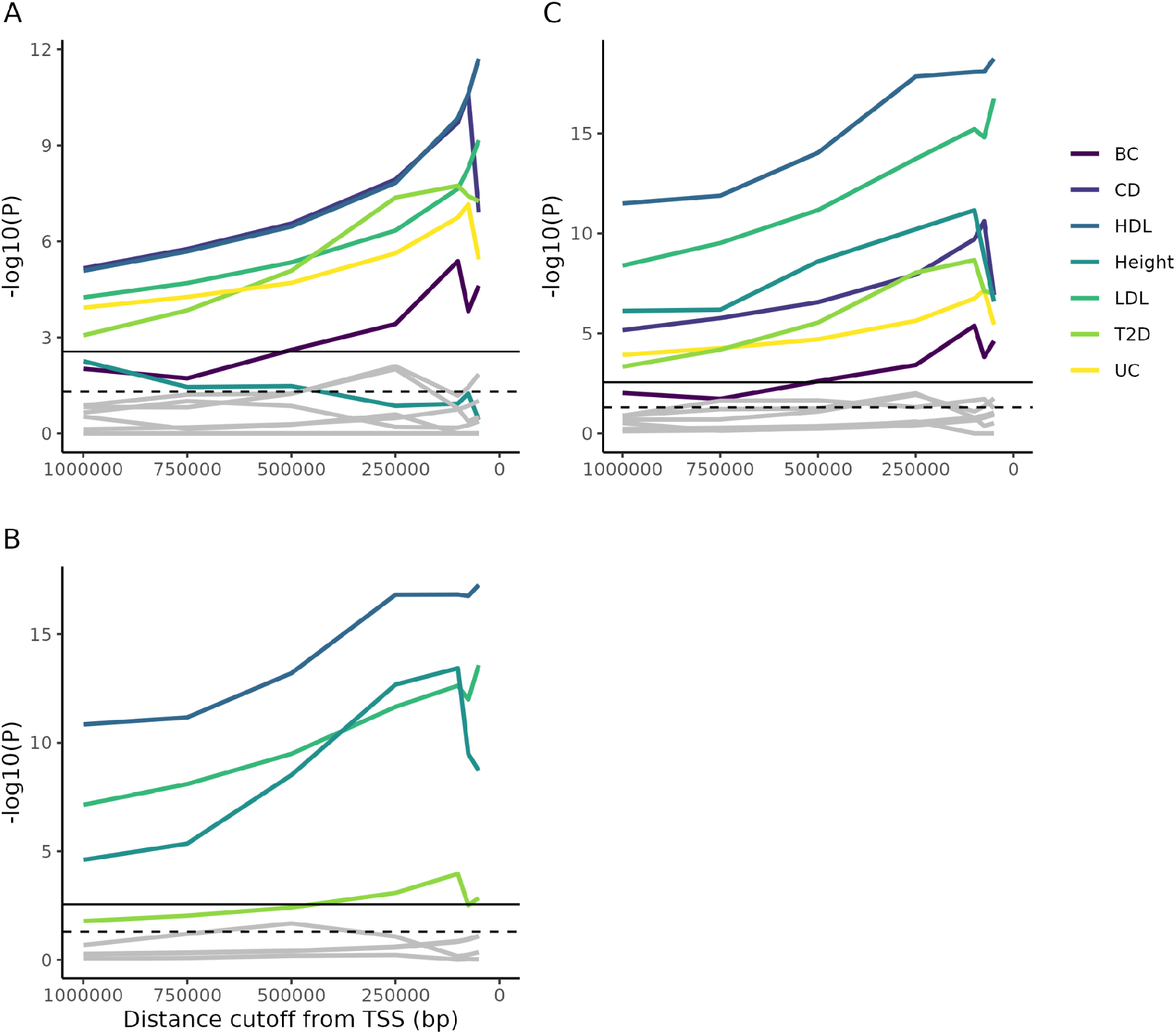
Enrichment of Mendelian genes near GWAS peaks. A) As the window around GWAS peaks shrinks, the enrichment of Mendelian genes within the window becomes increasingly significant, while the enrichment of non-matching trait pairs used as controls (gray lines; **Methods**) is not consistently increased. Some controls achieve nominal significance (dotted horizontal line), but none reach significance once multiple-testing is corrected for (solid horizontal line). B) As above, but for genes from Backman et al. (2021)^16^. C) The combined gene lists from parts A and B. Note that, accounting for multiple test correction (based on the total number of tests across all panels) height does not reach significance using the Mendelian gene list, while T2D is barely significant using the Backman list. However, combining the lists increases power and demonstrates significance for all traits.

**Supplementary figure 2.**
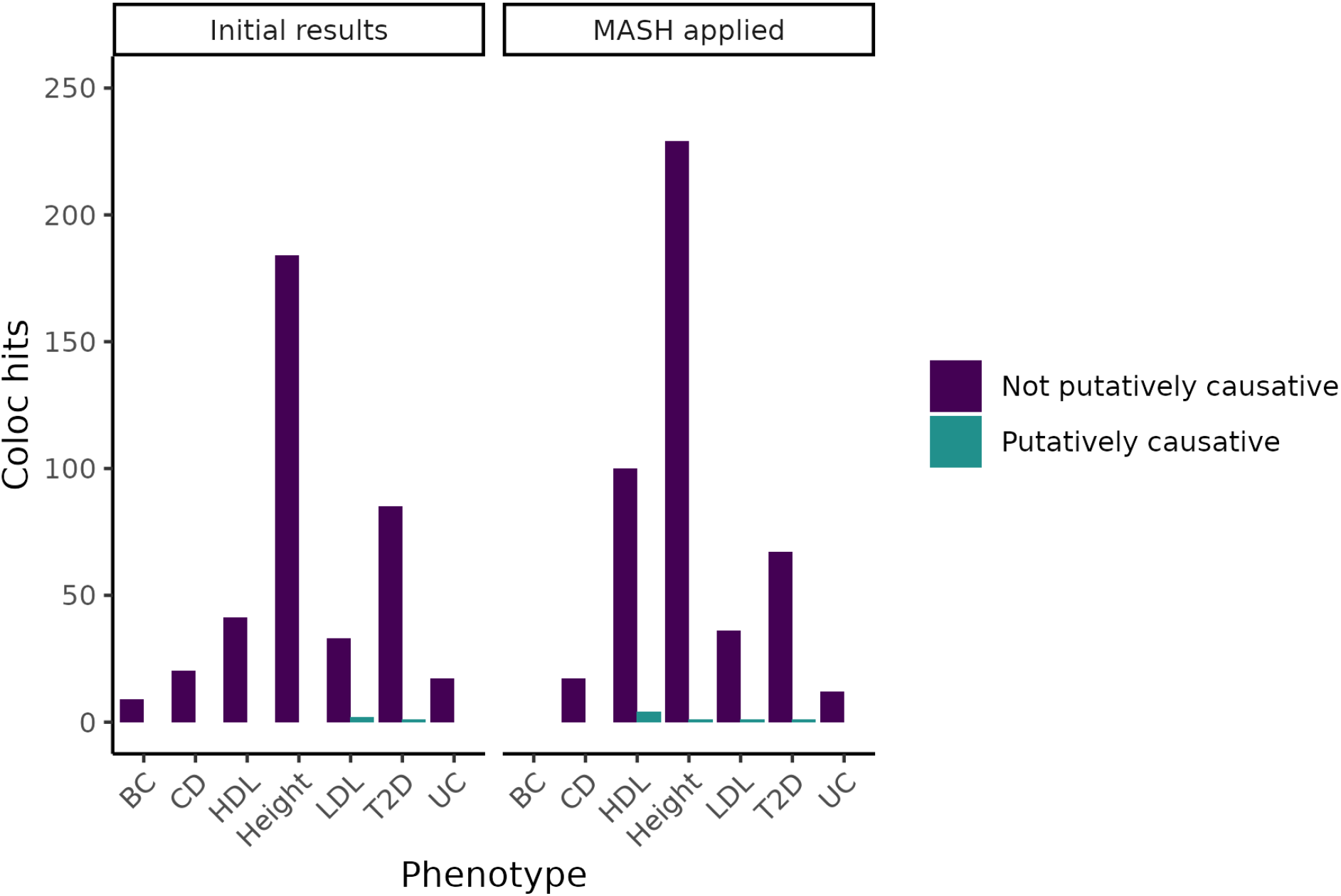
Change in coloc hits when adjusting eQTL statistics using MASH. By using the Bayesian method MASH to update our measurements of eQTLs based on tissues with similar expression patterns, we increased the number of colocalizations found. However, even in tissues in which the number of genes identified increased substantially, we did not meaningfully increase the number of putatively causative genes identified.

**Supplementary Table 1.** Roadmap epigenomics aliases of tissue types used for functional genomic analysis. Tissue types from the Roadmap Epigenomics Consortium do not perfectly match those from GTEx. However, there is overlap, and as with GTEx, we analyzed trait-relevant tissues.

| Alias | Descriptive Name |
| --- | --- |
| E027 | Breast myoepithelial primary Cells |
| E028 | Breast luminal epithelial Cells / Human mammary epithelial cells |
| E062 | Primary mononuclear cells from peripheral blood |
| E063 | Adipose nuclei |
| E066 | Liver |
| E075 | Colonic mucosa |
| E076 | Colon smooth muscle |
| E087 | Pancreatic islets |
| E095 | Heart left ventricle |
| E096 | Lung |
| E104 | Heart right atrium |
| E107 | Skeletal Muscle Male |
| E108 | Skeletal Muscle Female |
| E109 | Small Intestine |

**Supplementary Table 2.**
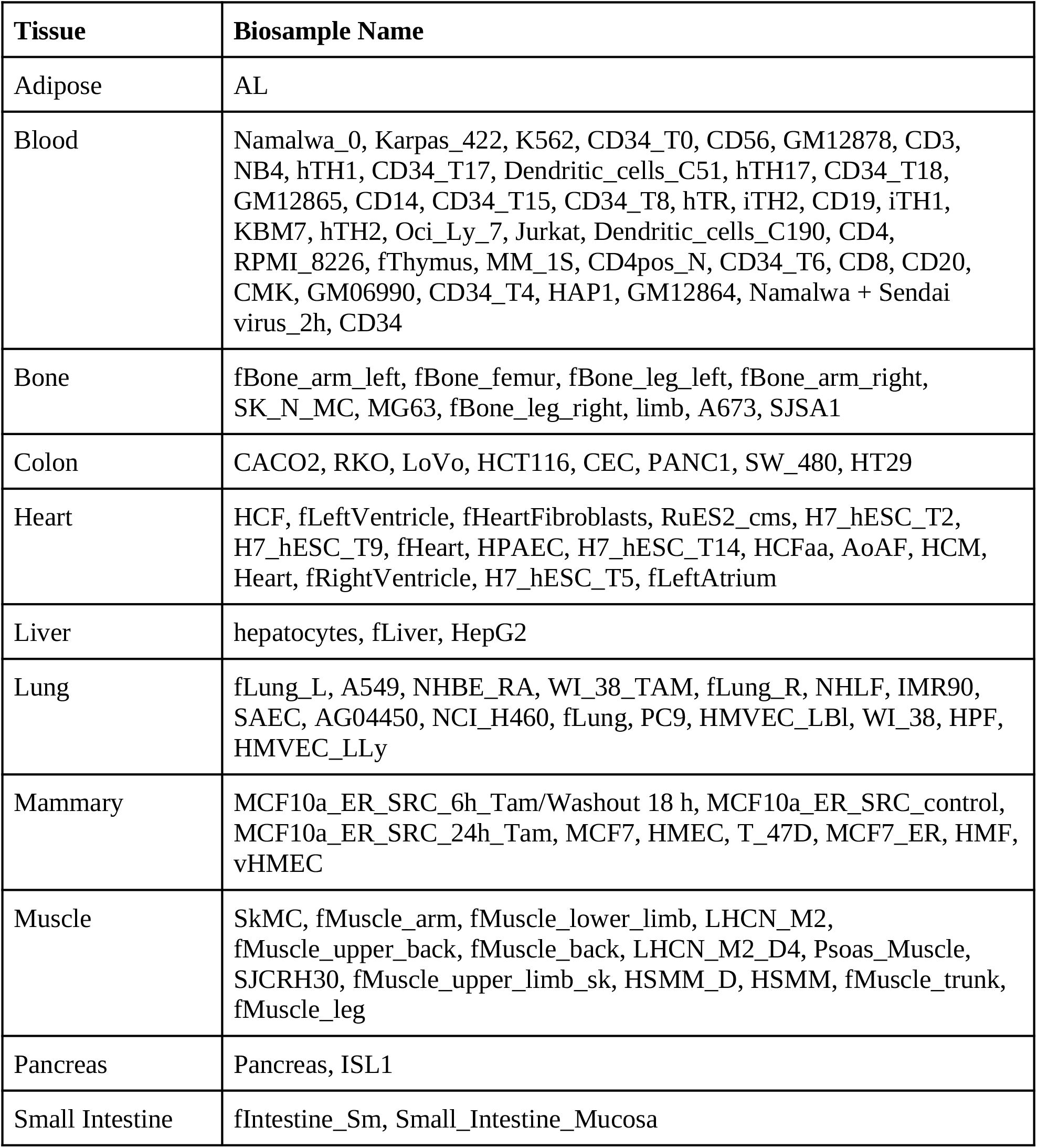
Tissue types and bio-samples from the DNase I hypersensitive sites index used for functional genomic analysis. Meuleman *et al*. assess DNase I hypersensitive sites across 438 cell and tissue types^39^; we selected the above based on their relevance to our complex traits.

## Supplementary Note 1. Evidence for the relationship between Mendelian and complex traits

More generally, this expectation is supported by several lines of evidence. Comorbidity between Mendelian and complex traits has been used to identify common variants associated with the complex traits^282^. Early GWAS found associations near genes identified through familial studies of severe disorders^283,284^, and later implicated some of the same genes in complex and Mendelian forms of cardiovascular^285^ and neuropsychiatric^286^ traits. More recent analyses have found that GWAS associations are enriched in regions near causative genes for cognate Mendelian traits in blood traits^287^, lipid traits and diabetes^15^, as well as a diverse collection of 62 traits^29^. Another recent method used transcriptomic, proteomic and epigenomic data to prioritize genes and found that, in a selection of nine phenotypes, selected genes were enriched for Mendelian genes causing similar traits^288^. Together, these suggest that genes causing Mendelian traits also influence cognate complex traits, but not through the same coding mechanism.

Genes can also harbor coding variants tied to less severe forms of a trait. These coding variants are more difficult to identify individually, as their effect sizes are much smaller. However, the greater number of variants (in aggregate) and freedom from searching for severe segregating traits, allows the use of large population datasets. Backman *et al*. used burden testing on UK Biobank data to identify genes whose coding variation affects complex traits, finding many genes not identified through familial studies^16^.

## Supplementary methods

### Gene selection

By manual literature search, we selected 128 genes harboring large-effect-size coding variants for one of the seven phenotypes (**Table 1**; specifically, we selected 128 gene-trait pairs, representing 121 unique genes). These genes were identified using familial studies, rare disease exome sequencing analyses, and, for breast cancer, using the MutPanning method^218^ (citations for each gene are included in **Table 1**). Review papers, as well as the OMIM database^289^, were generally used as starting points, but an examination of the original literature was needed to confirm genes’ suitability. For example, though SMC3 is known to cause Cornelia de Lange syndrome, which is characterized in part by short stature, SMC3 mutations lead to a milder form of the syndrome, usually without a marked reduction in stature^290^. Several of these phenotypes —height, HDL, cholesterol, breast cancer, and type II diabetes—were also analyzed in *Backman et al*., which, through burden testing, identified a total of 110 genes; after accounting for overlaps, this increased our set of putatively causative genes to 220^16^. The inclusion of genes from Backman *et al*. ensures that our results are not dependent on an undetected bias in our selection. The set of genes chosen from familial studies offered the advantage that it was selected based on independent methods and data distinct from the large-scale genotyping studies that have characterized the GWAS era. The tradeoff to this was the impossibility of selecting genes through a fully systematic and non-arbitrary process. Because this work was performed in the UKBB, there is some overlap between their data and ours. However, our work did not use exomes, and most of the variants driving their findings are too rare to influence GWAS results. When this is not the case, our decision to condition on coding variants should make the effects used in our work independent from their findings.

### Identifying coding variants

Because GWAS sample sizes are large enough to detect the low-frequency coding variants used to select some of our genes, it is possible that a coding SNP would distort the association signal of nearby eQTLs. To minimize this concern, we removed the effects of coding variants on GWAS. Many variants can fall within coding sequences in rare splice variants, so it is important to remove only those variants that appear commonly as coding. These coding SNPs were selected based on the pext (proportion of expression across transcripts) data^291^. Two filters were used. First, we removed genes whose expression in a trait-relevant tissue was below 50% of their maximum expression across tissues. Second, we removed variants that fell within the coding sequence of less than 25% of splice isoforms in that tissue. The remaining variants were used to correct GWAS signal, as explained below.

### GWAS

For height, LDL cholesterol, and HDL cholesterol, GWAS were performed using genotypic and phenotypic data from the UKBB. In order to avoid confounding, we restricted our sample to the 337K unrelated individuals with genetically determined British ancestry identified by Bycroft *et al*.^292^ The GWAS were run using Plink 2.0^293^, with the covariates age, sex, BMI (for LDL and HDL only), 10 principal components, and coding SNPs.

### Conditional analysis

Because UKBB has limited power for breast cancer, Crohn disease, ulcerative colitis, and type II diabetes, we used publicly available summary statistics. The Conditional and Joint Analysis (COJO)^27,28^ program can condition summary statistics on selected variants—in our case, coding variants—by using an LD reference panel. For this reference, we used TOPMed subjects of European ancestry^294^. The ancestry of these subjects was confirmed with FastPCA^295,296^ and the relevant data were extracted using bcftools^297^. Our conditional GWAS data are available at doi:10.5061/dryad.612jm644q

### Enrichment analysis

At each distance, the number of Mendelian and non-Mendelian genes within that window around GWAS peaks are counted. *P*-values are calculated using Fisher’s exact test (Supp. Fig. 1). Because Mendelian genes may be unusually important beyond our chosen traits, we conduct a set of controls by measuring the enrichment of non-matching Mendelian and complex traits (CD genes & BC GWAS; BC genes & LDL GWAS; LDL genes & UC GWAS; UC genes & height GWAS; height genes & T2D GWAS; T2D genes & HDL GWAS; HDL genes & CD GWAS).

### eQTL detection

eQTL summary statistics were taken from GTEx v7. Some methods detect colocalization with variants that are individually significant, but would not pass a genome-wide threshold^12^. Because we tested only a subset of genes and, we used the Bejamini-Hochberg method^298^ to calculate the FDR based on the number of tests we conducted multiplied by a correction factor to account for variants that are tested in combination with a gene but are not reported (a factor of 20 closely matched the genome-wide FDR results for GTEx). With this method 204/220 (93%) of our genes displayed an eQTL, including 134/147 genes with a nearby GWAS peak (91%). Even using the FDR statistics of the GTEx project—which are based on the assumption of testing every gene in every tissue—107/220 (49%) of our genes and 76/147 (52%) of genes near GWAS peaks had an eQTL at Q < 0.05.

### Colocalization

JLIM^12^ was run using GWAS summary statistics and GTEx v7 genotypes and phenotypes, for the tissues listed in **Table 2**. Coloc^11^ was run using GWAS and GTEx v7 summary statistics for the same tissues. eCAVIAR^14^ was run using GWAS and GTEx v7 summary statistics for these tissues, and a reference dataset of LD from UKBB^299^. MASH was run incorporating data from all non-brain tissues, and coloc was re-run using the adjusted values for the same tissues as before.

### MASH

Multivariate adaptive shrinkage (MASH) was applied to all GTEx tissues using the mashr R-package^32^. We restricted this model to non-brain tissues—which include all of our trait-selected tissues—due to the known tendency of brain and non-brait tissues to cluster separately in expression analysis^300–302^.

### Fusion (TWAS)

We used the FUSION implementation of TWAS, which accounts for the possibility of multiple *cis*-eQTLs linked to the trait-associated variant by jointly calling sets of genes predicted to include the causative gene, to interrogate our 220 loci^35^. FUSION included our putatively causative genes in the set identified as likely relevant to the GWAS peak in 66/220 loci (30%). However, interpretation of this TWAS result is difficult. For many complex traits, TWAS returns a large number of findings (e.g., over 150 for LDL cholesterol and over 4,800 for height). This is in part due to the multiple genes jointly returned at a locus, and can also be a result of the large number of tissues and cell types included in the implementation of FUSION. Most hits are found in tissues without any clear relevance to the trait, and absent in relevant tissues—LDL, for example, has more TWAS associations between expression and eQTL in prostate adenocarcinoma (24 genes associated), brain pre-frontal cortex (23 genes associated), and transformed fibroblasts (21 genes associated) than it does in adipose (16 genes associated), blood (11 genes associated), or liver (5 genes associated). Individual genes were often identified as hits in multiple tissues, but with an inconsistent direction of effect—that is, increased gene expression correlated with an increase in the quantitative trait or disease risk in some tissues, but a decrease in others, which suggests that the gene in question may not be the one whose expression contributes to the complex trait. Because of this possibility, and the known biological role of many of our genes, we restricted our results to tissues with established relevance to our traits.

### Fine-mapping GWAS hits

We fine-mapped the GWAS variants located within +/-100 kb of our putatively causative genes by applying the SuSiE algorithm^38^ on the unconditional summary statistics from the GWAS of breast cancer, Crohn disease, ulcerative colitis, type II diabetes, height, LDL cholesterol, and HDL cholesterol. An LD reference panel from UKBB subjects of European ancestry was used for this analysis. Fine-mapped variants were annotated using snpEff (v4.3t). Only non-coding variants were kept for further analysis.

### Functional genomic annotation of fine-mapped hits

We projected fine-mapped GWAS variants onto active regions of the genome, identified using three alternative approaches: (i) histone modification features, (ii) DNase I hypersensitive sites, and (iii) ChromHMM enhancers.

First, we looked at three histone modification marks, namely, acetylation of histone H3 lysine 27 residues (H3K27ac), mono-methylation of histone H3 lysine 4 residues (H3K4me1), and tri-methylation of lysine 4 residues (H3K4me3) from the Roadmap Epigenomics Project^37^ to identify functional enhancers which are key contributors of tissue-specific gene regulation. We downloaded imputed narrowPeak sets for H3K27ac, H3K4me1, and H3K4me3 from the Roadmap Epigenomics Project^37^ ftp site (https://egg2.wustl.edu/roadmap/data/byFileType/peaks/consolidatedImputed/narrowPeak/) for 14 different tissue types (**Supp. Table 1**). For each tissue type, we extracted the narrow peaks that are within +/-5 Mb of our putatively causative genes. Then following the approach described in Fulco et al.^36^, we extended the 150 bp narrow peaks by 175 bp on both sides to arrive at candidate features of 500 bp in length. All features mapping to blacklisted regions (https://sites.google.com/site/anshulkundaje/projects/blacklists) were removed. Remaining features were re-centered around the peak and overlapping features were merged to give the final set of features per histone modification track. Mean activity/strength of a feature (*A*_*F*_) was calculated by taking the geometric mean of the corresponding peak strengths from H3K27ac, H3K4me1, and H3K4me3 marks. We then combined these activity measurements with the linear distances between the features and the transcription start sites of causative genes to compute “activity-by-distance” scores (a simplified version of ABC scores^36^) for gene-feature pairs using the following formula.

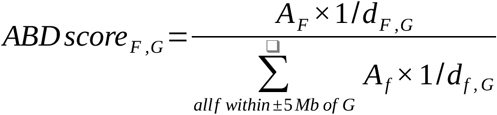

The ABD score can be thought of as a measure of the contribution of a feature, F to the combined regulatory signals acting on gene, G. A high ABD score may serve as a proxy for an increased specificity between a chromatin feature and the gene of interest. We projected the fine-mapped variants onto the chromatin features in different tissue types to assess whether there is an enrichment of likely causal GWAS hits in regulatory features near our putatively causative genes. Both proximity (genomic distance) and specificity (ABD scores) were considered to determine the regulatory contribution of the fine-mapped hits.

Next, we looked at the DNase I hypersensitive sites (DHSs) which are considered to be generic markers of the regulatory DNA and can contain genetic variations associated with traits and diseases^39^. We downloaded the index of human DHS along with biosample metadata from https://www.meuleman.org/research/dhsindex/. The index was in hg38 coordinates which were converted to hg19 coordinates using the online version of the hgLiftOver package (https://genome.ucsc.edu/cgi-bin/hgLiftOver). We created a DHS index for each tissue type relevant to the traits and diseases we analyzed by including all DHS that are present in at least one bio-sample from a certain tissue type (**Supp. Table 2**). We then selected DHS that lie within +/-100 kb of the TSS of our putatively causative genes. Since DHS are of variable widths, we recentered the summits in a 350 bp window and merged overlapping sites in the same way as we did for other chromatin marks. We calculated the mean activity (*A*_*F*_) by averaging the strengths of all the merged sites. Next, we calculated the activity by distance score for each DHS and gene pair using the same formula described above. Finally, for each fine-mapped SNP, we identified all DHS that fall within +/-100 kb of the SNP.

Finally, we used in-silico chromatin state predictions (chromHMM core 15-state model^37^) for relevant tissue types (**Supp. Table 1**) to identify active enhancer regions in the genome. Tissue-specific chromHMM annotations were downloaded from the Roadmap Epigenomics Project^37^ ftp site (https://egg2.wustl.edu/roadmap/data/byFileType/chromhmmSegmentations/ChmmModels/coreMarks/jointModel/final/). We considered a fine-mapped variant to fall in an enhancer region if it was housed within a chromHMM segment described as either *enhancer*, or *bivalent enhancer*, or *genic enhancer*. Since chromHMM annotations are not accompanied by activity measurements, the ABD approach could not be applied here.

## Acknowledgements

We thank Alkes Price, Alex Bloemendal, Benjamin Neale, Bogdan Pasanuic, Sasha (Alexander) Gusev, and Matt Warman for their helpful discussions. This research was supported by NIH grants R35GM127131, R01HG010372, R01MH101244, and U01HG012009,. N.J.C was supported by NIH training grant T32GM74897. UK Biobank was accessed under projects 14048 and 10438. TOPMed data were used under dbGaP project 28674.

